# Designing welfare-maximising financing for publicly-provisioned digital child-health platforms: A mixed-methods policy simulation from Thailand’s KhunLook application

**DOI:** 10.64898/2026.04.26.26351784

**Authors:** Kiatanantha Lounkaew

**Affiliations:** Faculty of Economics, Thammasat University, Bangkok, Thailand

**Keywords:** digital health, willingness to pay, welfare simulation, means-tested subsidy, maternal and child health, health financing, Thailand

## Abstract

National digital health platforms are scaling faster than the evidence on how to finance them. This paper develops a welfare-simulation framework that converts a published willingness-to-pay (WTP) distribution into a prescriptive pricing recommendation, applied to Thailand’s KhunLook maternal-and-child-health application. Predicted WTP values at the 25th, 50th and 75th unconditional quantiles and the OLS mean — drawn from a survey of n = 680 Thai parents and relatives of young children previously reported in Lounkaew et al. (2025) — enter the simulation as parametric inputs. Quintile-level WTP is imputed by monotone-cubic interpolation, a population of 250,000 caregivers is drawn from truncated-Normal distributions around the quintile means, and five financing scenarios are compared: full public provision (S1), a flat market-priced fee (S2), freemium (S3), fine-grained income-tiered pricing (S4), and a means-tested subsidy with a flat fee for the top 60% (S5). A thematic reading of Thai digital-health policy documents bounds the institutionally feasible scenario set and anchors the interpretation of the simulation numbers. Full public provision maximises total welfare at 437.4 million THB but runs a five-year fiscal deficit. The means-tested subsidy gives up about 15% of that welfare to recover 198.6 million THB in net producer surplus, distributes consumer surplus toward lower-income quintiles (concentration index −0.258), and plugs into the existing Thai state welfare card register at near-zero marginal administrative cost. The ranking holds across all twelve sensitivity specifications. Administrative simplicity in subsidy targeting, read against the Thai WTP distribution, dominates finer-grained tiering on both welfare and equity grounds. The framework transfers cleanly to other middle-income countries deciding how to price a national digital health platform.

**Author summary:** Many middle-income-country governments now run free national smartphone apps for the health of mothers and young children, but the funding model is increasingly fragile as initial donor and research grants run out. The question this paper asks is simple: if such a platform had to start charging, what pricing structure would raise the most money without locking out the families who need the app most? Using a published Thai survey of 680 parents and relatives of young children, the paper simulates five alternative designs — free, flat fee, freemium, fine-tiered by income quintile, and a means-tested subsidy — and finds that offering the bottom 40% of households free access while charging the top 60% a flat 395 Thai baht per year (roughly USD 11) captures 85% of the welfare of the status-quo free model, generates 199 million baht of fiscal surplus over five years, and distributes benefits toward lower-income users rather than toward the well-off. The design works because Thailand’s state welfare card register already identifies the low-income target population, so means-testing is essentially free to administer. Other countries with comparable social registries can apply the same logic to their own digital health platforms.

## Introduction

Governments across the low- and middle-income world have stood up national digital health platforms for mothers and young children at a pace that has outstripped the evidence on how to keep them running. The WHO gave the trajectory an institutional stamp in 2019 [3], and a growing body of evaluation work has documented user engagement, behavioural change and social returns from these platforms in settings as varied as Karachi, rural India, urban and rural Kenya, Tanzania’s Kibaha district, Italian paediatric day-hospitals, and UK early-years services [1,2,4,23,25,26,27]. Less has been said about financing. Once donor and grant funding ends — a recurrent concern across LMIC digital-health programmes in sub-Saharan Africa and South Asia [5,29,31], and not least in Thailand where KhunLook has cycled through several rounds of research-council support [6] — how should a ministry of health structure cost recovery without walking back the equity commitments that made the platform worth building in the first place?

Three financing models are in active use. A platform can be fully public: a ministry absorbs implementation and maintenance costs from general revenue, and users pay nothing. It can be priced at market, with households paying directly or through insurance. Or it can sit in between — freemium, tiered, or means-tested — using price discrimination to extract willingness-to-pay (WTP) from higher-income users while keeping access open for the poor. Evaluation work has been thorough on the first step of valuation — estimating what users are willing to pay [4,5] — but rarely converts that valuation into a specific pricing rule when equity, uptake and fiscal sustainability all have to be satisfied at once. The present paper takes that second step.

Thailand and the KhunLook application are a useful case. The application was launched in 2013 by a multi-institutional team with support from the Health Systems Research Institute, the Ministry of Public Health and UNICEF [4,6], and has passed 550,000 downloads across iOS and Android.

It is the explicit digital successor to the Maternal and Child Health Handbook — the Pink Book — which the Department of Health has distributed free since 1985 and which, after a 1997 amendment to the Civil Registration Act, serves as the statutory vehicle for birth-certificate issuance [7,8]. Few other LMIC digital child-health platforms carry that kind of institutional lineage. The lineage is also what makes the financing question awkward: continuing to distribute KhunLook free is faithful to the Pink Book precedent but keeps the platform on the public fiscal purse; charging for it crosses a boundary the Thai health system has been careful about.

A financing recommendation that ignores institutional realism is not useful. Welfare simulations that treat pricing as an abstract exercise — maximise surplus subject to a budget constraint — can yield designs that no Thai planner could actually administer. Fine-grained income-based pricing is the clearest illustration: the Thai health system has no infrastructure for identifying a user’s income quintile at the point of service, and building that infrastructure for the sake of one app would be hard to cost-justify. A mixed-methods approach that pairs the welfare arithmetic with a thematic reading of the relevant policy documents [9] is better placed to produce recommendations that can be implemented as well as defended.

The question at the centre of this paper is which of five financing designs for a publicly-provisioned digital child-health platform best balances welfare, equity and fiscal sustainability in a middle-income setting. The coefficient vector from Lounkaew et al. [4] supplies the parametric inputs; a thematic reading of Thai digital-health policy documents constrains which scenarios are worth simulating and how to read their results; the welfare simulation does the arithmetic. The contribution is prescriptive policy design rather than descriptive valuation, and the framework is intended to transfer to other LMIC digital-health financing decisions.

## Materials and Methods

### Study context and the KhunLook platform

KhunLook is a mobile application developed in Thailand in 2013 by a partnership of academic researchers, the Ministry of Public Health and UNICEF, and distributed at no cost on iOS and Android [4,6]. At the time of the underlying user survey the application had been downloaded more than 550,000 times. A conservative cumulative download base of 500,000 is used in the simulation that follows, matching the planning figures in the Integrated Evaluation report [4] and avoiding attribution of benefits to post-survey downloads. Functionality covers growth and developmental monitoring, vaccination reminders, longitudinal health records from birth through age six, and in-app guidance from paediatric and dental professionals [6]. The application is the digital successor to Thailand’s Maternal and Child Health Handbook (the Pink Book), in continuous use since 1985 and still the primary physical record in antenatal and well-child services [7].

The platform is currently free at the point of use and funded from the public purse, with residual support from international partners. The policy question is not whether to price KhunLook but how the financing architecture should be designed to maximise population welfare given the heterogeneous willingness to pay for digital child-health services across Thai households, and given the implementation constraints of a middle-income universal health coverage system [10].

### Study design

This study is a secondary analysis. The quantitative inputs are drawn from a survey of n = 680 Thai parents and relatives of young children — the primary user base of KhunLook — commissioned and funded by the Health Systems Research Institute (HSRI) of Thailand and reported in Lounkaew et al. [4]; no new data were collected and no coefficients were re-estimated.

The HSRI user survey provides a cross-sectional, quantitative willingness-to-pay (WTP) distribution, whose published OLS and unconditional-quantile coefficient estimates (reproduced here as Table 1) are used as parametric inputs to the welfare simulation. The qualitative component is a separate corpus — Thai digital-health policy documents, legislative instruments and historical accounts of the Maternal and Child Health Handbook — analysed by inductive thematic analysis following Braun and Clarke [9]. The qualitative work does two jobs: it constrains which of several imaginable financing scenarios are worth simulating in the first place, and it interprets the simulation numbers against the institutional realities that determine whether a given scenario is actually implementable.

**Table 1.** Willingness-to-pay regression results from Lounkaew et al. [4].

| Variable | OLS | UQR q25 | UQR q50 | UQR q75 |
| --- | --- | --- | --- | --- |
| Constant | -43.667***<br>(3.258) | 20.244***<br>(4.312) | -6.126*<br>(3.568) | 17.561***<br>(0.345) |
| Log income | 8.598***<br>(0.605) | 4.539***<br>(0.790) | 2.078***<br>(0.655) | -2.384***<br>(0.064) |
| (Log income)<sup>2</sup> | -0.372***<br>(0.028) | -0.196***<br>(0.036) | -0.083***<br>(0.030) | 0.127***<br>(0.029) |
| Years of Schooling | 0.000<br>(0.000) | 0.039***<br>(0.009) | 0.016**<br>(0.007) | -0.001<br>(0.006) |
| (Years of Schooling)<sup>2</sup> | 0.013<br>(0.009) | -0.001<br>(0.000) | -0.001<br>(0.000) | 0.000<br>(0.000) |
| Age | -0.017<br>(0.022) | -0.017<br>(0.017) | -0.036**<br>(0.015) | -0.023<br>(0.020) |
| Age<sup>2</sup> | 0.000<br>(0.000) | 0.000<br>(0.000) | 0.000<br>(0.000) | 0.000<br>(0.000) |
| Occupation | 0.025<br>(0.019) | 0.066***<br>(0.016) | 0.046***<br>(0.015) | 0.035**<br>(0.017) |
| Number of Children | 0.058***<br>(0.013) | 0.014<br>(0.012) | 0.003<br>(0.011) | 0.003<br>(0.011) |
| Family size | 0.008<br>(0.005) | 0.001<br>(0.004) | 0.001<br>(0.004) | 0.003<br>(0.004) |
| Gender | 0.026<br>(0.019) | -0.011<br>(0.016) | 0.016<br>(0.015) | 0.027*<br>(0.016) |
| Single / Widowed | 0.007<br>(0.018) | 0.031**<br>(0.015) | 0.016<br>(0.015) | -0.004<br>(0.016) |
| Apartment | 0.023<br>(0.018) | 0.000<br>(0.015) | -0.010<br>(0.015) | -0.007<br>(0.016) |
| Townhouse | 0.003<br>(0.018) | 0.025*<br>(0.015) | 0.022<br>(0.015) | -0.010<br>(0.016) |
| Stand-alone house | -0.006<br>(0.019) | 0.003<br>(0.016) | -0.018<br>(0.015) | -0.014<br>(0.016) |
| Renting property | 0.044**<br>(0.019) | -0.008<br>(0.015) | 0.001<br>(0.015) | 0.017<br>(0.016) |
| **R<sup>2</sup> / Pseudo-R<sup>2</sup>** | **0.711** | **0.516** | **0.387** | **0.313** |
| **N** | **680** | **680** | **680** | **680** |

Note. Dependent variable is the natural logarithm of annual willingness-to-pay for KhunLook (in THB). Coefficients reported above standard errors (in parentheses). Significance: \*\*\* $p < 0.01$ ; \*\* $p < 0.05$ ; \* $p < 0.10$ , computed as $|\beta/SE|$ against Normal critical values. Sample size $n = 680$ in all specifications. OLS reports $R^2$ ; UQR specifications report Pseudo- $R^2$ following [11]. Source: Lounkaew et al. [4] Table 6.

### Source of the willingness-to-pay distribution

Lounkaew et al. [4] estimated WTP through a log-linear model of the form

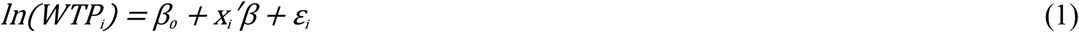

where x_i_ is a vector of caregiver covariates (log household income and its square, years of schooling and its square, age and its square, indicators for occupation, number of children, family size, gender, marital status, housing type and rental status), and ε_i_ is a mean-zero disturbance term. The coefficient vector β was estimated by ordinary least squares (OLS) and separately by unconditional quantile regression (UQR) following Firpo, Fortin and Lemieux [11]. UQR proceeds in two stages. First, the dependent variable is transformed through the Recentered Influence Function,

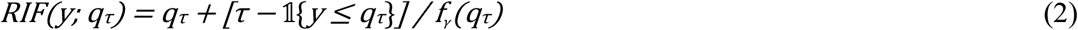

where qτ is the τ-th unconditional quantile of WTP and f_γ_(qτ) is the density at that quantile. Second, the transformed variable is regressed on the covariates by OLS to obtain quantile-specific marginal effects β(τ). Table 1 reproduces the full estimated coefficient vector for OLS and for τ ∈{0.25, 0.50, 0.75} with standard errors and conventional significance indicators.

Three patterns in Table 1 matter for what follows. The income terms do most of the work: log income and its square are strongly significant across all four specifications, with signs indicating diminishing marginal valuation up to the median and a convex reversal at the 75th quantile. Any pricing rule keyed to income needs to respect that non-linearity. Occupation (a mid- to high-skilled dummy) is significant at all three quantiles but not at the mean — a distributional signal that a conditional-mean regression misses. Most demographic and housing controls do not clear conventional significance thresholds, which is why the simulation below concentrates on income-quintile variation as the dominant predictable source of WTP heterogeneity.

The predicted WTP values used in the simulation are the point predictions from these coefficients evaluated at sample means: 341.0 Thai baht (THB) at the OLS mean, and 342.3, 395.3 and 441.3 THB at τ = 0.25, 0.50 and 0.75 respectively. Descriptive statistics for the underlying WTP sample are mean = 364.22 THB, standard deviation = 97.29 THB, and range [40, 589] THB. Implementation cost is 9.521 million THB and the active user base is 250,000, equal to a 50% activation rate on 500,000 cumulative downloads [4]. The simulation is exactly consistent with Table 1 at the predicted WTP anchors and at the R^2^ statistic that drives the within-quintile dispersion; no coefficient in the table is re-estimated.

### Quintile-level WTP imputation

Individual-level income-quintile WTP values are not published in [4]. Quintile-mean WTP is imputed by fitting a monotone piecewise-cubic Hermite interpolating polynomial (PCHIP) through five anchor percentile points — (0, 40), (25, 342.3), (50, 395.3), (75, 441.3), (100, 589) in (percentile, THB) space — and evaluating the interpolated inverse cumulative distribution function at the income-quintile midpoints p_k_ ∈{10, 30, 50, 70, 90}:

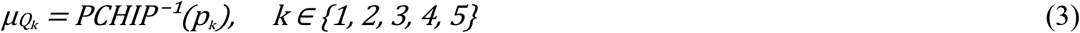

yielding μ_{Q1} = 199.2, μ_{Q2} = 357.8, μ_{Q3} = 395.3, μ_{Q4} = 429.1, and μ_{Q5} = 515.2 THB (Figure 1, Table 3). The recovered population mean is 378.1 THB, within 3.8% of the descriptive mean 364.22 THB. Within-quintile residual dispersion is set at

**Fig 1.**
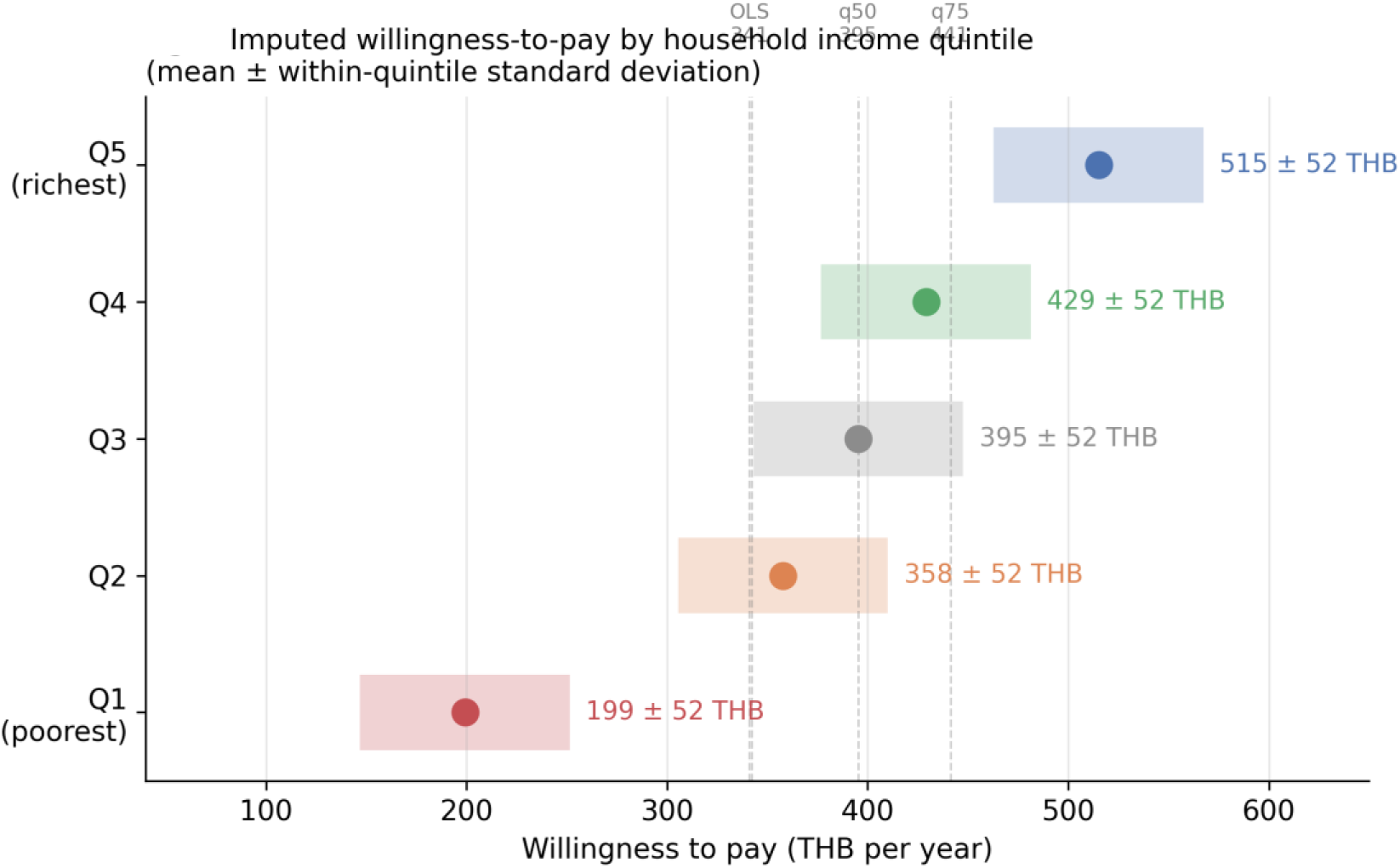
Imputed willingness-to-pay by household income quintile. Mean willingness-to-pay with 1-standard-deviation within-quintile dispersion. Dashed vertical lines mark published OLS and UQR anchor points. Source: author calculations from Lounkaew et al. [4].

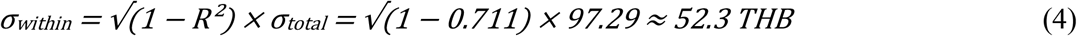

with the OLS R^2^ taken directly from Table 1. Each caregiver i in quintile Qk is assigned a simulated WTP from a truncated Normal distribution:

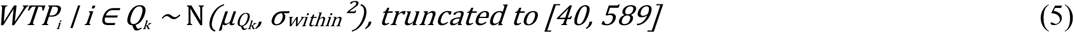

The simulation draws M = 50,000 caregivers per quintile, giving a population of 250,000 that matches the empirical active user base. The random seed is fixed at 42 for reproducibility.

### Qualitative data and method

The qualitative corpus is entirely secondary and is drawn from Thai government and international policy documents. It comprises the National Digital Health Strategy [12], the 11th and 12th National Health Development Plans [13,14], the Civil Registration Act provisions that integrated the birth certificate into the Pink Book [8], documentation of the UNICEF-supported ‘9 Yang Pua Srang Luk’ (9 Steps to Build Your Child) programme [15], and historical accounts of the Maternal and Child Health Handbook since 1985 [7]. The HSRI research report [4] is cited in this paper only as the source of the quantitative WTP coefficients reproduced in Table 1; its narrative chapters are not treated as qualitative data. No new interviews or primary qualitative data were collected.

Coding followed Braun and Clarke’s six steps [9]: familiarisation, inductive open coding, collation into candidate themes, review against the source material, definition and naming, and selection of extracts. Coding was conducted by a single coder (the author). Trustworthiness is addressed through the codebook in Supporting Information S1, triangulation across the three distinct source sets, and reliance on publicly citable material. Inter-coder reliability is not reported — a limitation discussed below.

### Welfare-simulation framework

Under each financing scenario s, a simulated caregiver i adopts the platform if and only if her simulated WTP is at least as large as the price she faces under s:

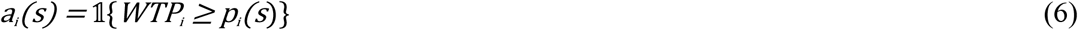

where p_i_(s), defined by the pricing rule for scenario s, may depend on i’s income quintile (Table 2). Individual consumer surplus conditional on adoption is

**Table 2.** Financing scenarios and pricing rules.

| Scenario | Pricing rule (THB/yr) | Policy rationale |
| --- | --- | --- |
| S1 | 0 for all users; fully public | Status quo; maximum-uptake benchmark with zero revenue |
| S2 | Flat fee 341 (OLS-mean WTP) | Naive market-based benchmark; quantifies welfare cost of excluding low-WTP households |
| S3 | Free basic tier; premium at 441 (q75) | Freemium two-tier common in DHIs; tests willingness to pay for depth |
| S4 | Tiered: Q1=0; Q2=342; Q3–Q4=395; Q5=441 | Ability-to-pay design aligned with Thai UHC equity principles |
| S5 | Q1–Q2=0; Q3–Q5=395 (q50) | Means-tested subsidy mirroring Thai state welfare card eligibility |

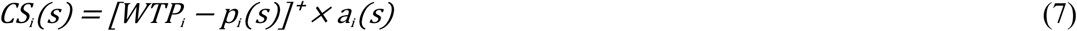

with [·]^+^ = max(·, 0). Scaling the simulated population of M caregivers up to the N-caregiver user base and discounting over the T-year horizon at social discount rate r gives the aggregate monetary measures

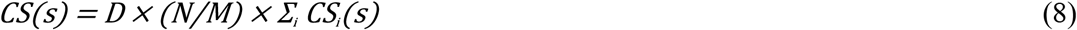

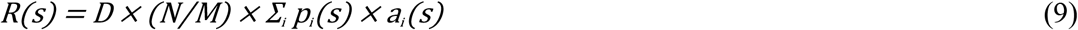

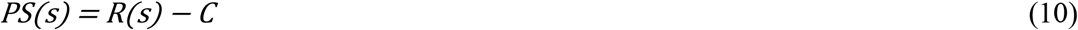

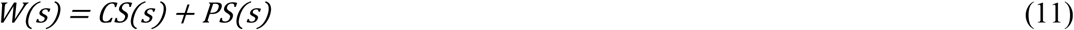

where the annuity discount factor is

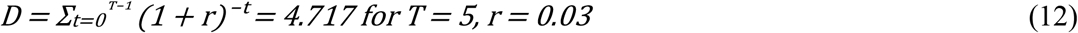

and C is the up-front implementation cost (9.521 million THB). Deadweight loss among non-adopters, on the assumption that the marginal cost of serving an additional user of a non-rival digital service is zero, is

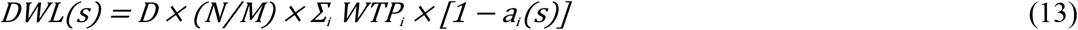

Equity outcomes are the Q1-to-Q5 access gap and the Wagstaff-type concentration index of consumer surplus over income rank [17]:

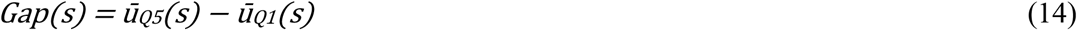

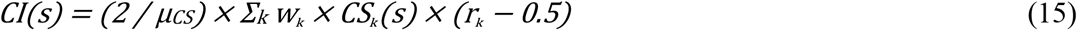

where ū_Q1 and ū_Q5 are the uptake shares in quintiles 1 and 5, w_K_ = 1/5 is the population share of quintile k, CS_K_(s) is the mean consumer surplus in quintile k under s, r_K_ is the fractional-rank midpoint of quintile k (0.1, 0.3, 0.5, 0.7, 0.9), and μ_{CS} is the population-mean consumer surplus. Negative values of CI indicate pro-poor distribution of surplus; positive values indicate pro-rich capture. Total welfare is reported alongside the access gap and the concentration index rather than collapsed into a single welfare-plus-equity scalar — the trade-off is the interesting object.

### Financing scenarios

The five scenarios in Table 2 are chosen to span the realistic Thai policy space rather than to exhaust the theoretical one. For S1, S2, S4 and S5 the pricing rule is a direct function of the individual’s income quintile; S3 (freemium) needs its own two-tier decomposition, set out after the table.

In the freemium case (S3), a caregiver gets value V_basic from the free basic tier and an incremental value from purchasing the premium tier at price p_prem. The decomposition is parameterised by a share α ∈(0, 1) governing how much of the caregiver’s total WTP is captured by the basic tier alone:

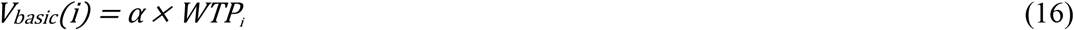

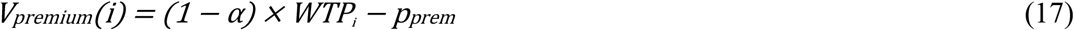

with premium adoption indicator a_i_^p^ = ᵽ9{(1 − α) × WTP_i_ ≥ p_prem}. The base-case value is α = 0.60 — three-fifths of the platform’s perceived value coming from its basic functions (growth monitoring, vaccination reminders, milestone tracking), two-fifths from a hypothetical premium tier such as direct specialist consultation, expanded developmental content, or personalised nutrition guidance. The sensitivity analysis varies α over {0.50, 0.60, 0.70}.

### Sensitivity analysis

Twelve sensitivity specifications were run. Implementation cost C was varied by ±25% (7.14 to 11.90 million THB). The social discount rate r was tested at 0%, 3% (base case) and 5%; the 3% baseline follows the Thai Health Technology Assessment guideline convention for public-sector investment evaluation [16]. The effective user base N was tested at 150,000, 250,000 and 500,000. Scenario prices were varied by ±20%. The freemium share parameter α was varied as above. As a robustness check on the Marshallian consumer-surplus base case, equivalent variation was computed under a Cobb-Douglas preference approximation,

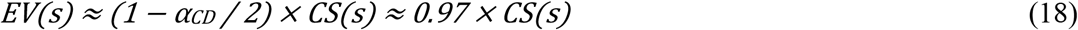

with α_CD = 0.06 as a conservative share-of-budget parameter for digital health services. An extreme-case run combined the lowest cost and highest user base assumptions to test the joint envelope. Full tables are in Supporting Information S2.

## Ethics statement

This paper analyses secondary coefficient estimates from Lounkaew et al. [4] and publicly available policy documents. No primary data collection was conducted. The survey data that produced the WTP estimates were collected under the ethics framework documented in the Integrated Evaluation report for the Health Systems Research Institute of Thailand. No individual-level data are accessed here. Ethics review was not required.

## Results

Results proceed in four stages. The qualitative themes come first because they shape how the quantitative numbers should be read. The imputed quintile-level WTP distribution follows. Scenario-level welfare outcomes and the welfare–equity frontier come third. Sensitivity and the integrated findings close the section.

### Qualitative themes shaping financing feasibility

The thematic analysis produced six themes (full codebook in Supporting Information S1). Theme 1 — Historical continuity from Pink Book to KhunLook. The Department of Health has distributed the Pink Book free since 1985 [7], and since 1997 the book has served as the statutory vehicle for birth-certificate issuance [8]. KhunLook was positioned as the digital continuation of that line rather than as a market-based alternative [4,6]. The practical implication is that any pricing regime at odds with the free-at-point-of-use public-good tradition will face political and institutional resistance that does not show up in a welfare calculation.

Theme 2 — Equity as a constitutive value of Thai universal health coverage. The Universal Coverage Scheme introduced in 2002 embedded access equity as a foundational design principle [10]. In the policy corpus, departures from free-at-point-of-use delivery are the exception that needs justifying, not the rule. Scenarios producing access gaps between income quintiles are politically disfavoured regardless of how they perform on aggregate efficiency. Theme 3 — Digital literacy as an uptake constraint. Several sources flagged uneven digital capability among lower-income and rural caregivers as a binding constraint on uptake that is independent of price [3,18].

Eliminating price as a barrier does not automatically produce universal adoption; a design that assumes it will is vulnerable to overstating its own welfare gains.

Theme 4 — The public-private boundary in Thai health financing. Thai health policy maintains a clear conceptual boundary between goods that belong in the public sphere and goods that households buy privately [12,19]. The corpus treats digital child-health monitoring as public-sphere, consistent with the Pink Book precedent. A freemium design that asks households to pay for premium digital features crosses that boundary — policy actors will resist unless the basic tier is demonstrably non-inferior. Theme 5 — Sustainability of donor-seeded digital innovations. Thai policy documents carry explicit concerns about the durability of digital-health initiatives dependent on international donor funding [5,6,20]. The KhunLook financing question is inseparable from whether the platform can become fiscally self-sustaining within the Thai public budget.

Theme 6 — Administrative capacity for means-testing. Thailand runs a state welfare card (Bat Sawatdikan Haeng Rat) whose eligibility criteria target low-income households and approximate the bottom two income quintiles [21]. The administrative machinery for identifying and subsidising that population therefore already exists and is reusable. Fine-grained quintile-specific pricing (S4 below) would require capacity the Thai health system does not presently have; coarse subsidy targeting (S5) can be implemented through the existing welfare-card register at near-zero marginal administrative cost.

### Imputed WTP distribution by income quintile

The imputed distribution (Figure 1, Table 3) is sharply unequal. Mean WTP in the bottom quintile (199.2 THB) is less than 40% of the mean in the top quintile (515.2 THB). That gradient mechanically drives everything that follows: any pricing rule ignoring it translates into an access gap between Q1 and Q5.

**Table 3.** Imputed quintile-level WTP distribution used in simulation.

| Quintile | Mean WTP (THB/yr) | Within-Q SD (THB) | Interpretation | Share of sample |
| --- | --- | --- | --- | --- |
| Q1 (poorest) | 199.2 | 52.3 | Price-sensitive | 20% |
| Q2 | 357.8 | 52.3 | Near population mean | 20% |
| Q3 | 395.3 | 52.3 | Median | 20% |
| Q4 | 429.1 | 52.3 | Upper-middle | 20% |
| Q5 (richest) | 515.2 | 52.3 | High-value buyer | 20% |

*Note.* Quintile-mean WTPs imputed via PCHIP monotone-cubic interpolation through the percentile anchors implied by Table 1 predictions. Within-quintile SD derived as $\sqrt{(1-R^2)} \times \sigma$ from OLS $R^2 = 0.711$ (Table 1, column 1) and overall $\sigma = 97.29$ THB.

### Scenario-level welfare outcomes

Table 4 carries the headline numbers. All monetary quantities are in millions of THB, discounted over five years at 3%.

**Table 4.** Welfare outcomes by financing scenario.

| Scenario | Uptake | CS | Revenue | PS | W | DWL | Gap | CI |
| --- | --- | --- | --- | --- | --- | --- | --- | --- |
| S1 Free (status quo) | 100.0% | 446.9 | 0.0 | −9.5 | **437.4** | 0.0 | 0.00 | +0.148 |
| S2 Flat fee 341 THB | 68.7% | 82.6 | 276.1 | 266.6 | 349.2 | 88.2 | +1.00 | +0.460 |
| S3 Freemium | 100% / 0% premium | 268.1 | 0.0 | −9.5 | 258.6 | 0.0 | 0.00 | +0.148 |
| S4 Income-tiered | 75.7% | 86.4 | 261.8 | 252.2 | 338.6 | 98.7 | −0.08 | −0.260 |
| **S5 Subsidy Q1–Q2 + flat 395 Q3–Q5** | **84.7%** | **174.2** | **208.2** | **198.6** | **372.9** | **64.5** | **−0.01** | **−0.258** |

*Note.* CS = consumer surplus; PS = net producer surplus; W = total welfare; DWL = deadweight loss; Gap = Q5 uptake minus Q1 uptake; CI = Wagstaff-type concentration index of consumer surplus (negative = pro-poor). All monetary quantities in millions of Thai baht, discounted over 5 years at 3%. S5 (bold) is the recommended design.

### Integrated findings

Table 4 and Figure 2, read together, bring four patterns into focus. S1 peaks on total welfare at 437.4 million THB, but that peak is bought with a persistent 9.5-million-THB fiscal deficit — the implementation cost is never recovered. The naive flat fee at the OLS-mean WTP, S2, shuts Q1 out of the platform entirely and produces the only strongly pro-rich concentration index in the table (+0.460). That is a quantitative illustration of what the policy corpus describes narratively: untargeted market pricing builds rather than narrows the digital divide [1,3,22]. S3 is revealing for a different reason. At the base-case α = 0.60, no quintile’s residual premium valuation (1 − α) × WTP exceeds the 441-THB premium price, premium uptake collapses to zero, and freemium pricing at q75 is simply not viable in this distribution. S4, the fine-grained income-tiered design that policy intuition tends to favour, generates the largest deadweight loss among the priced designs (98.7 million THB) because Q3 and Q4 face tier prices close to their own mean WTP and partially exit the market.

**Fig 2.**
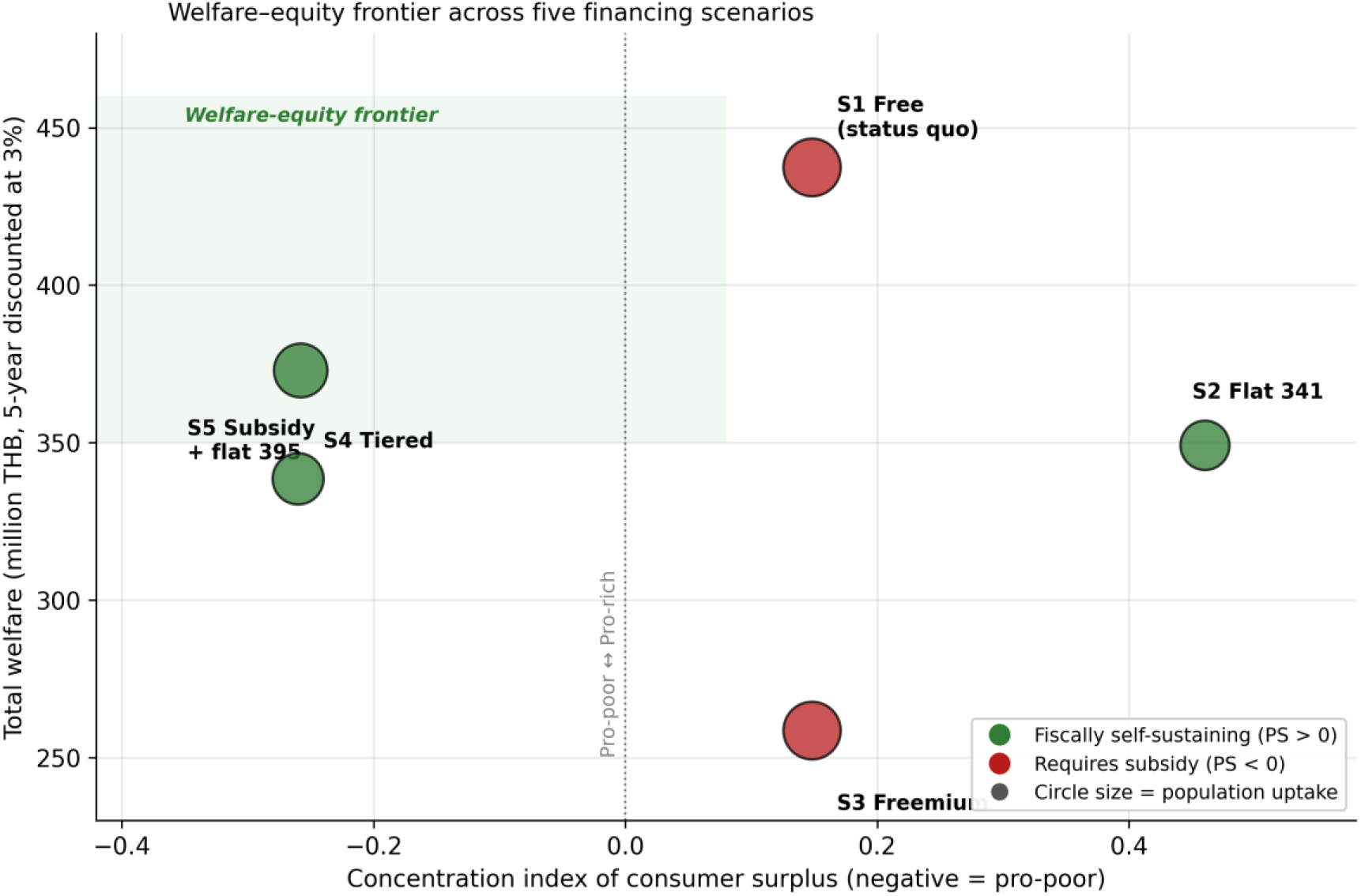
Welfare-equity frontier across the five financing scenarios. Circle size proportional to population uptake; colour indicates fiscal sustainability (green = PS > 0; red = PS < 0). Shaded region marks the welfare–equity frontier (high welfare, pro-poor capture). S5 occupies the frontier; S1 achieves higher welfare but with fiscal deficit; S2 is strongly pro-rich.

S5 is the scenario that dominates the welfare–equity frontier. Its total welfare (372.9 million THB) is the highest among fiscally self-sustaining designs, within 15% of the S1 maximum. Net producer surplus over the five-year horizon reaches 198.6 million THB — enough to fund further investment in the platform at more than twenty times the baseline implementation cost. The concentration index of −0.258 distributes consumer surplus toward lower-income quintiles. S5 dominates S4 because pooling Q3 through Q5 at the median WTP price (395 THB) eliminates the middle-quintile exclusion that S4 creates by pricing tiers close to each quintile’s own mean. Administrative simplicity in subsidy targeting, under the Thai WTP distribution, is not a second-best compromise: it wins on welfare and equity simultaneously. Theme 6 reinforces the recommendation from the institutional side. Thailand’s state welfare card register [21] already identifies the low-income target population, so implementing S5 is close to costless administratively; implementing S4, by contrast, would require quintile-identification capacity the Thai health system does not currently have.

### Sensitivity analysis

The ranking is stable across all twelve sensitivity specifications. S1 remains the welfare-maximum in 12 of 12 runs; S5 remains the welfare-maximum among fiscally self-sustaining designs in 12 of 12 runs. The one parameter to which the results are visibly sensitive is the scenario price. At −20% price, S4 and S5 converge on total welfare (428.5 vs 431.8 million THB) because lowering the Q3–Q4 tier price eliminates middle-quintile exclusion. At +20% price, S4 collapses to W = 137.2 million THB as Q3, Q4 and part of Q5 are priced out, while S5 remains robust. S3 welfare rises to 321.8 million THB at α = 0.50 and falls to 201.4 million THB at α = 0.70; no tested value of α pushes S3 above either S1 or S5. Full tables are in Supporting Information S2.

## Discussion

The question at the centre of this paper — how to finance a publicly-provisioned digital child-health platform in a middle-income country where willingness-to-pay rises steeply with income — has a specific answer conditional on the Thai data and policy context. A means-tested subsidy that gives the bottom 40% of households free access and charges the top 60% a flat 395 Thai baht per year captures 85% of the welfare frontier set by the fully-public status quo, runs a 198.6-million-THB fiscal surplus over five years, distributes consumer surplus in a pro-poor direction (CI = −0.258), and keeps that ranking across every one of the twelve sensitivity specifications.

The contribution is of three kinds. On method, the paper pushes welfare-economic evaluation of digital public goods past descriptive valuation and into prescriptive design [11,17]. Recent empirical work in digital health — Ahmer and colleagues’ Karachi case study [1], the DEEP cognitive-assessment line from proof-of-concept in rural Haryana [24] to longitudinal non-specialist-delivered validation [2], the MomCare value-based programme in urban and rural Kenya [26], the Afya-Tek digital referral platform linking public and private primary care in Tanzania [27], and the WHO’s 2019 guideline [3] — has been strong at measuring value and weaker at translating value into price. The simulation framework here closes part of that gap. On substance, the finding that coarse means-testing beats fine-grained tiering runs against a pricing-policy intuition that finer calibration should extract more surplus without harming access. The intuition breaks down because fine tiers place the middle of the WTP distribution dangerously close to its own price, turning would-be adopters into priced-out non-adopters. On context, the paper adds specificity to an LMIC digital-health literature that has documented digital divides at length [3,22,29,31] but rarely named the financing instruments that could close or widen them.

The S5 recommendation survives an institutional-feasibility check against the qualitative themes, which is the point of the mixed-methods design. Keeping the basic platform free preserves the Pink Book public-good tradition (Theme 1) and the universal-coverage principle in Thai UHC (Theme 2); means-testing only the upper tier keeps the financing inside the Thai public sphere rather than pushing core services into private payment (Theme 4); the producer surplus it generates addresses the sustainability concern that has haunted donor-seeded Thai digital-health initiatives (Theme 5); and the existing state welfare card register handles eligibility without building new administrative capacity (Theme 6). The theme S5 does not fully resolve is Theme 3. Eliminating the price barrier is necessary but not sufficient for universal uptake among the bottom two quintiles, because digital-literacy gaps bind independently of price — fewer than one in three older or less-literate Nepali adults can effectively use mobile health tools even where access is free [31], and stakeholders consulted on England’s mandated 2–2½-year child-development review warned that digitisation without complementary in-person support deepens rather than narrows exclusion [28]. Without a complementary investment in community health worker outreach, caregiver digital-skills training, or in-clinic onboarding at antenatal visits, the welfare calculus here will overstate what is realisable in practice.

Four caveats shape how the recommendation should be read. First, the WTP estimates that feed the simulation inherit the limitations of the underlying stated-preference instrument: hypothetical bias, social-desirability bias in an online convenience sample, and no formal control group for validity testing [4]. Second, the quintile-level WTP distribution used here is imputed from published aggregate statistics rather than fit to individual-level data; the imputation recovers the population mean to within 3.8%, but tail values in Q1 and Q5 depend on the distributional assumption and would be tighter if fit to the source dataset directly. Third, the freemium share parameter α is an assumption rather than an estimate; no value in the tested range {0.50, 0.60, 0.70} overturns the scenario ranking, but readers who prefer α outside that window will want to re-run the S3 column. Fourth, the welfare framework is static: it abstracts from network externalities (which would make S1 and S5 look better as adoption scales), from platform-cost economies of scale, and from the dynamic gains from preventive-care behavioural change documented in the published behavioural outcomes. Absolute welfare numbers are therefore likely conservative; rankings should be robust. The framework also abstracts from the opposite side of digital exposure: evidence that unregulated child and adolescent screen use is itself a growing public-health concern [30] suggests that welfare gains from any child-health platform should be weighed against the broader screen-exposure baseline of its intended users, and the platform designed to be bounded and purposeful rather than simply accessible. A single-coder qualitative analysis is a fifth caveat; the codebook (Supporting Information S1) is transparent and the sources citeable, but a second coder and a kappa statistic would be a standard upgrade for a revised version.

The Thailand-specific implication is concrete. The National Digital Health Strategy [12] and the ‘9 Yang Pua Srang Luk’ (9 Steps to Build Your Child) programme [15] both commit to scaling KhunLook as a national child-health platform. S5 is a financing pathway consistent with those commitments that reduces the platform’s dependence on general revenue and on international donor cycles. The Health Systems Research Institute, which funded the underlying WTP study [6], is well placed to pilot S5 against the existing free-for-all model in a single province — using the state welfare card register [21] for eligibility, and the PromptPay national payment rail for collecting fees from higher-income users — before any national rollout. That pilot would also be the natural test of the simulated uptake and welfare numbers against real-world behaviour.

The LMIC-generalisable version of the argument is more cautious but follows the same logic. Recent implementation-science frameworks for scaling digital health in LMIC settings — the digital fit/viability model proposed for sub-Saharan Africa [29] and the infrastructure–policy– literacy triad argued for Nepal [31] — treat economic viability, a financing rule that outlives the donor cycle, as a distinct and often binding condition alongside individual, technical, task, organisational and legal fit. In any middle-income country where (i) household WTP for digital child-health services rises steeply with income, (ii) an existing means-testing register identifies the bottom two quintiles for other social-protection purposes, and (iii) the public-health system treats maternal and child health as within the public sphere rather than the private market, a means-tested subsidy design with a flat fee for higher-income users should dominate both fine-grained tiering and flat market pricing. Prices and costs will differ, but the structural result — administrative simplicity in targeting beats precision when a usable register already exists — is transferable. Countries without a usable register face a genuinely different problem, for which full public provision may remain the only practical recommendation.

Willingness-to-pay data, once they exist, can do more than describe. Treated as an input to policy design and interpreted through the institutional constraints of the delivery system, they point to a specific financing rule. For Thailand’s KhunLook platform, that rule is a means-tested subsidy: the bottom 40% of households get free access and the top 60% pay a flat 395 Thai baht per year. The design captures 85% of the welfare frontier set by the status quo, generates 198.6 million THB of fiscal surplus over five years, distributes consumer surplus toward lower-income users, and runs on Thailand’s existing state welfare card infrastructure at negligible marginal cost. The ranking holds across every sensitivity specification tested. The broader point is methodological. Welfare simulation paired with institutional thematic analysis produces pricing recommendations that can be defended analytically and implemented practically. The combination is portable to other middle-income digital-health platforms facing the same question.

## Data Availability

The datasets used and analysed during the current study are available at the discretion of the Health Systems Research Institute (HSRI), Thailand.

## Acknowledgments

I am the sole author.

## Ethical approval and consent to participate

Not applicable. The empirical analysis is based on secondary data from the Health Systems Research Institute (HSRI), Thailand; no new primary data were collected for this study.

